# Prevalence and Factors Associated With Delay in Presentation of Breast Cancer Patients in Ethiopia: A Cross-Sectional Institution-Based Study

**DOI:** 10.1101/2022.11.01.22281792

**Authors:** Jabir Abdella Muhammed, Eric Sven Kroeber, Bedilu Deribe, Susanne Unverzagt, Lesley Taylor, Amdehiwot Aynalem, Deriba fetene

**Affiliations:** School of Nursing, Collage of medicine and health sciences, Wachemo University, Hosanna, SNNPR, Ethiopia; Center of Health Sciences, Institute of General practice, Martin-Luther-University Halle-Wittenberg, Halle, Sachsen-Anhalt, Germany; School of nursing, Collage of medicine and health sciences, Hawassa University, Hawassa, Sidama regional state, Ethiopia; City of Hope National Medical Center, Duarte, CA, USA; Department of Nursing, Collage of medicine and health science, Madawalabu University, Robe, Ethiopia

**Keywords:** Breast cancer, delayed presentation, Hawassa, Ethiopia, low-and middle-income countries, observational study

## Abstract

**Background:** In developing countries, the high mortality of breast cancer (BC) patients is strongly related to delayed presentation and subsequent advanced stage diagnosis, pointing to the need for improved detection programs. This study aims to assess the prevalence and factors associated with delayed presentation of BC patients at Hawassa University Oncology Center (HUOC), Hawassa, Ethiopia.

**Methods:** A cross-sectional institution-based survey was conducted among BC patients between May 1^st^ and August 30^th^, 2021. BC patients attending HUOC were included by consecutive sampling. Data was collected using an interviewer-administered questionnaire and medical record data extraction. A multivariable binary logistic regression model was carried out to identify associations between delayed presentation and potential risk factors.

**Results:** A total of 150 BC patients participated in the study giving a response rate of 100%. Of these, 86 (57.3%) women presented with a long delay of ≥ 3 months. The median time to visit a health care provider after recognition of the first symptoms was 5.5 months. Urban residence (adjusted odds ratio (AOR) = 0.42; 95 %-CI=0.18-0.97) and not visiting of traditional healer (AOR=0.15, 95 %-CI=0.07-0.34) was associated with shorter delay time. No breast pain symptoms (AOR=8.57; 95 %-CI=3.47-21.15), no family history of BC (AOR=5.12; 95 %-CI=1.36-19.33), and travel distance ≥ 5 km (AOR=3.00, 95 %-CI=1.25-7.21) were associated with longer delay in presentation.

**Conclusion:** A substantial proportion of patients with BC in southern Ethiopia have delays in seeking medical care. Interventional programs like public BC awareness campaigns are strongly needed to reduce delayed presentation and to increase early detection of cancer cases.

## Background

Breast cancer (BC) is one of most frequently diagnosed cancers and occurs mostly in women (1). Globally, BC is the leading cause of death with about 2.3 million new cases and 685,000 estimated deaths in 2020 (2). In Africa the BC mortality rate is the highest among the continents and the incidence rate is increasing (3). In Ethiopia, around 60,000 new cases of BC are diagnosed annually (4). According to the Addis Ababa cancer registry report data, BC accounts for 33% of all female cancer cases and 23% of all cancers (5). In developed countries, more than 70% of BC patients are diagnosed at an early stage and have good prognosis with an overall survival rate of 90% (6)(7). In low- and middle-income countries (LMIC) BC patients are frequently diagnosed at advanced stages with only 20 to 60% of the patients being diagnosed in early stages (8). Main reasons are poor community awareness as well as poor diagnostic and treatment services (9). Delayed presentation to the health facilities after recognition of first symptoms largely contributes to advanced stage diagnosis of BCs, resulting in significantly lower survival rate compared to women who seek medical attention on time (10). More than half (54%) of female BC patients in black African populations search medical care later than after three months after recognition of symptoms (11). Reports from other LMICs have found delays as high as 45.9% in Karachi, Pakistan (12) and 31.7% in Iran (13). In Ethiopia, available data ranged from 50.5% (North East Ethiopia) (14) of BC patient seeking health care with a delay of more than three months to 75.5% (Gondar town, North West Ethiopia) (15).Socio-demographic factors linked to delayed presentation of BC are advanced age (16)(17)(18), being divorced or widowed (19)(17), lower education level (20)(21)(15)(14), travel distance >5 km (17), living in rural areas (15)(14), initial uptake of traditional medicine (18)(22), negative history of family BC (23), previous history of benign breast disease, other co-morbidities (24) and limited knowledge on screening practices like breast self-examination (25) are clinical and health-related factors associated to delayed presentation. The Ethiopian national cancer control plan published in 2015 outlines an aspiring strategy for improving early diagnosis of BC by promoting awareness and breast examination for women coming to health institutions (26). However, the prevalence of and factors associated with patient delay for seeking medical care remain high in Southern parts of Ethiopia where strong information is lacking particularly (27). Moreover, the factors linked to patient delay in presentation vary from area to area depending on socio-cultural and health care system variety. Understanding the current obstacles for early diagnosis for BC patients is helpful for early management and treatment of patients with BCs leading to a better prognosis. Therefore, this study aims to assess the prevalence and factors associated with delay in presentation of BC patients at Hawassa university comprehensive specialized hospital (HUCSH) cancer treatment center, the only oncology unit in the Southern part of Ethiopia.

## Methods and Materials

### Study design, setting and population

An institution-based cross-sectional study was conducted among BC patients attending cancer treatment center in HUCSH, Hawassa, Ethiopia between May 1^st^ and August 30^th^, 2021. HUCSH is a teaching hospital in the Sidama region and the region’s only comprehensive specialized hospital, providing cancer treatment and management service for more than 18 million people living in the Sidama, southern nations and nationalities, and some of Oromia regions of Ethiopia. We consecutively included all female BC patients aged above 18 years who received a new pathologically confirmed diagnosis of BC during the data collection period. We excluded patients who were seriously ill and unable to communicate due to disabilities.

### Sample size and sampling procedure

Sample size calculation was done using Epi Info software using a 95% confidence interval (CI) and 5% margin of error and an assumed proportion of patients with delayed presentation of 50.5% (from a comparable study from Dessie referral hospital oncology center, Ethiopia (14)) and resulted in a sample size of n=137 patients. Assuming a non-responder-rate of approximately 10%, 150 patients were recruited.

### Data collection tools and procedure

We used an interviewer-administered questionnaire and a medical record data extraction tool. The structured questionnaire was adapted from the Dessie study (14) and contextualized local differences as well as differing research objectives. The tool was initially prepared in English then translated to the national language Amharic. The Amharic version was used for data collection. The tool comprised socio-demographic factor, clinical and health system variables. A pretest was conducted on 5% of the sample size at Tikur Anbessa specialized hospital, Addis Ababa, Ethiopia before initiation of the main study. Necessary corrections were made based on the results of the pretest. Data collection was done by two data collectors who have first degree in nursing and are familiar with the study area. The data collectors were trained by the principal investigator about data collection, the content of the questionnaire, objectives and relevance of the study, confidentiality of information, right and approach to participants. Face to face interviews were implemented among consenting women. Date of diagnosis and stage of disease information was documented from medical hospital records. Staging was done according to American joint committee on cancer (AJCC). To minimize recall bias, patients were encouraged to connect important time points of first recognition of breast abnormalities and first visit of healthcare with calendrically referenceable dates like holidays, new years and others. According to the Aarhus model (28), the health-seeking interval was defined between the date of first symptom recognition by the patient and the first date the patient visited a health care provider. Waiting for more than three months (≥ 90 days) before consulting a health care provider was considered as delayed presentation.

### Data management and analysis

The collected data was coded, entered, and cleaned using Epi-data software version 4.6 and exported to SPSS (Statistical Package for Social Sciences) version 25 for further analysis. Bivariate and multivariate analysis was carried out to investigate the association between the delayed presentation variable and potential risk factors. Those variables having p values less than 0.25 on bivariate logistic regression were entered into the multivariate logistic regression to control all possible confounders and identify associations. The result of the final model was expressed in terms of adjusted odds ratio (AOR) with 95%CI. P-value <0.05 was considered as statistically significant.

### Ethical Consideration

Ethical clearance was obtained from Hawassa University College of medicine and health science, institutional review board. A formal letter of cooperation to conduct this research was written to HUCSH cancer treatment center. Verbal and written consent was obtained from each respondent.

## Results

### Socio-demographic Characteristics of study participants

A total of 150 BC patients participated in the study giving a response rate of 100%. The age of the participants ranged from 25 to 65 years with the mean age being 37.4 years. Of all participants, 41.3% were unable to read and write, 39.3% were housewife or jobless, 52.7% lived in rural community, and 66% were married. The average monthly income was 3500 Ethiopian Birr (ETB) ($71.75) with a range of 500 to 9000 ETB ($10.25-$184.5). Ninety percent of the participants had already given birth, 78% of the participants reported that it takes more than 5km from their home to the Hospital oncology center (Table 1).

**Table 1:** socio-demographic characteristics of the study population.

| Socio-demographic variables | Frequency (n=150) | Percent (%) |
| --- | --- | --- |
| Age |  |  |
| < 35 years | 66 | 44 |
| ≥ 35 years | 84 | 56 |
| Educational status |  |  |
| Unable to read and write | 62 | 41.3 |
| Read and write | 36 | 24 |
| Primary education | 23 | 15.3 |
| Secondary education | 14 | 9.3 |
| Collage and above | 15 | 10 |
| Occupation |  |  |
| Housewife or jobless | 59 | 39.3 |
| Governmental | 55 | 36.7 |
| Merchant | 22 | 14.7 |
| Farmer | 14 | 9.3 |
| Residence |  |  |
| Urban | 71 | 47.3 |
| Rural | 79 | 52.7 |
| Marital status |  |  |
| Married | 99 | 66.0 |
| Single, widowed, or divorced | 51 | 34.0 |
| Monthly income in ETB |  |  |
| < 1500 | 46 | 30.7 |
| 1500-4500 | 54 | 36.0 |
| > 4500 | 50 | 33.3 |
| Gave birth |  |  |
| Yes | 135 | 90.0 |
| No | 15 | 10.0 |
| Parity |  |  |
| $\leq 3$ | 88 | 58.7 |
| 3-5 | 38 | 25.3 |
| > 5 | 24 | 16.0 |
| Travel distance |  |  |
| < 5 km | 33 | 22.0 |
| $\geq 5$ km | 117 | 78.0 |
| Time to come to HUCSH |  |  |
| less than 6hour | 42 | 28.0 |
| 6 to 12 hours | 59 | 39.3 |
| 12 or more hours | 49 | 32.7 |

Abbreviations for the table: ETB, Ethiopian birr. HUCSH, Hawassa university comprehensive specialized hospital

### Clinical characteristics of the study population

More than half of the participants (55.3%; n=83) had never heard about BC before their illness. Almost two thirds (63.3%) reported that a breast lump was the first presenting symptom, 48.7% reported breast pain, and 41% discharge. Eighty-six participants (57.3%)initially considered their symptoms to be nothing serious, and 52.7% visited a traditional healer before seeking health care. More than half of the participants (57.3%) did not perform breast self-examination. Thirty-four participants (22.7%) had a known family history of BC. The most common reasons to seek a healthcare provider were persistence of the symptom (n=97; 64.7%) and recommendation of another person (n=30; 20%) (Table 2).

**Table 2:** clinical characteristics of women with breast cancer at the oncology center of HUCSH, 2021.

| **Characteristics** | **Frequency (n=150)** | **Percent (%)** |
| --- | --- | --- |
| Have you Heard of breast cancer previously |  |  |
| Yes | 67 | 44.7 |
| No | 83 | 55.3 |
| **The first change of breast** |  |  |
| Breast lump |  |  |
| Yes | 95 | 63.3 |
| No | 55 | 36.7 |
| Breast pain |  |  |
| Yes | 73 | 48.7 |
| No | 77 | 51.3 |
| Swelling of the breast |  |  |
| Yes | 24 | 16.0 |
| No | 126 | 84.0 |
| Discharge |  |  |
| Yes | 61 | 40.7 |
| No | 89 | 59.3 |
| Ulceration of the breast |  |  |
| Yes | 48 | 32.0 |
| No | 102 | 68.0 |
| Dimpling of breast |  |  |
| Yes | 49 | 32.7 |
| No | 101 | 67.3 |
| Pulling in of the nipple |  |  |
| Yes | 61 | 40.7 |
| No | 89 | 59.3 |
| Your first interpretation of the symptoms |  |  |
| Nothing serious | 86 | 57.3 |
| Infection | 48 | 32.0 |
| Cancer | 16 | 10.7 |
| Visited traditional healer before seeking health care |  |  |
| Yes | 79 | 52.7 |
| No | 71 | 47.3 |
| Breast self-examination |  |  |
| Yes | 64 | 42.7 |
| No | 86 | 57.3 |
| Family history of breast cancer |  |  |
| Yes | 34 | 22.7 |
| No | 116 | 77.3 |
| The reason that made you visit the health care provider |  |  |
| Knowledge of breast cancer symptom | 23 | 15.3 |
| Persistence of the symptom | 97 | 64.7 |
| Recommendation of another person | 30 | 20.0 |

### Prevalence of delay in presentation of breast cancer patients

Out of 150 BC patients who participated in this study, 57.3 % (n=86; 95 % CI=49-65.4%) women were delayed for ≥ 90 days (≥3 months). The mean delay time was 5.5 months. Among the delayed women 73 (84.9%) were diagnosed at a late stage (Table 3). The odds ratio for a late stage in case of patient delay was 8.2 with a 95% CI of 3.8 to 17.8. The most common reasons for delay in presentation were belief that it will relief by itself (n=91; 60.7%), use of traditional treatment option (n=79; 52.7%) and lack of money for medical care and transport (n=71; 47.2%).

**Table 3:** stages of breast cancer at diagnosis of patients attending HUCSH cancer treatment center.

| Stages of the disease at diagnosis | Patient delay |  |
| --- | --- | --- |
|  | < 90 days | ≥ 90days |
| Early Stage (I&II) | 38 | 13 |
| Late stage (III&IV) | 26 | 73 |

### Factors associated with delayed presentation of breast cancer patients

In the multivariate logistic regression analysis, home residence, breast pain, family history of BC, visit of traditional healer and travel distance were significantly associated with delay in presentation. Patients with an urban residence were 58.5% less likely to delay than rural residents (AOR=0.41; 95 %-CI 0.18 to 0.97). Similarly, BC patients who had no breast pain were approximately nine times more likely to delay than their counterparts (AOR=8.57; 95%CI=3.473-21.153). In addition, women without a family history of BC were five times more likely to delay (AOR=5.12; 95%CI=1.356-19.329). BC patients who have not visited traditional healer were 84.8% less likely to delay than who visited traditional healers (AOR=0.152, 95%CI=0.068-0.344). Moreover, those patients who travel ≥ 5km from home to the healthcare provider were about three times more likely to delay (AOR=2.997, 95%CI=1.247-7.206) (Table 4).

**Table 4:**
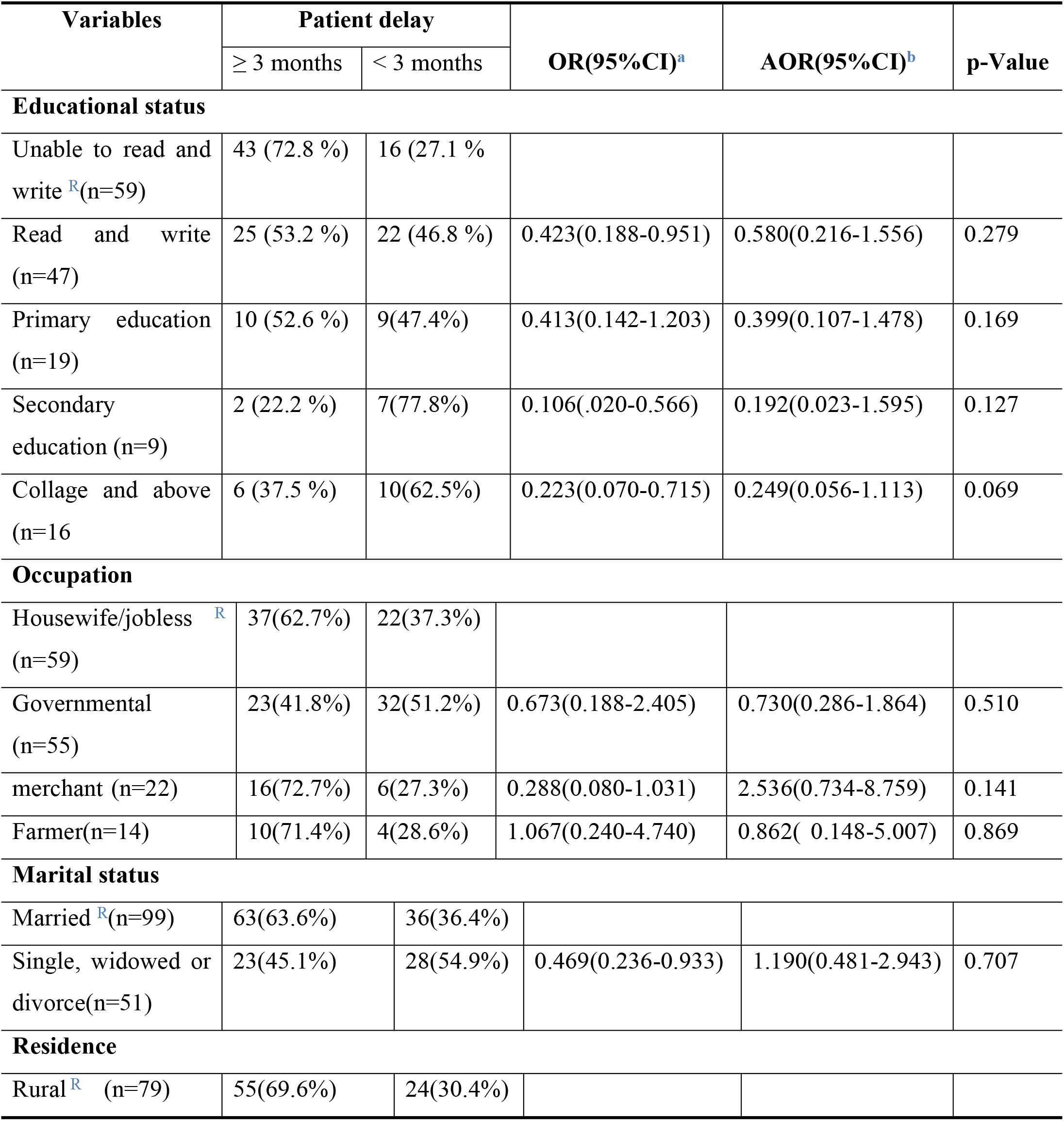
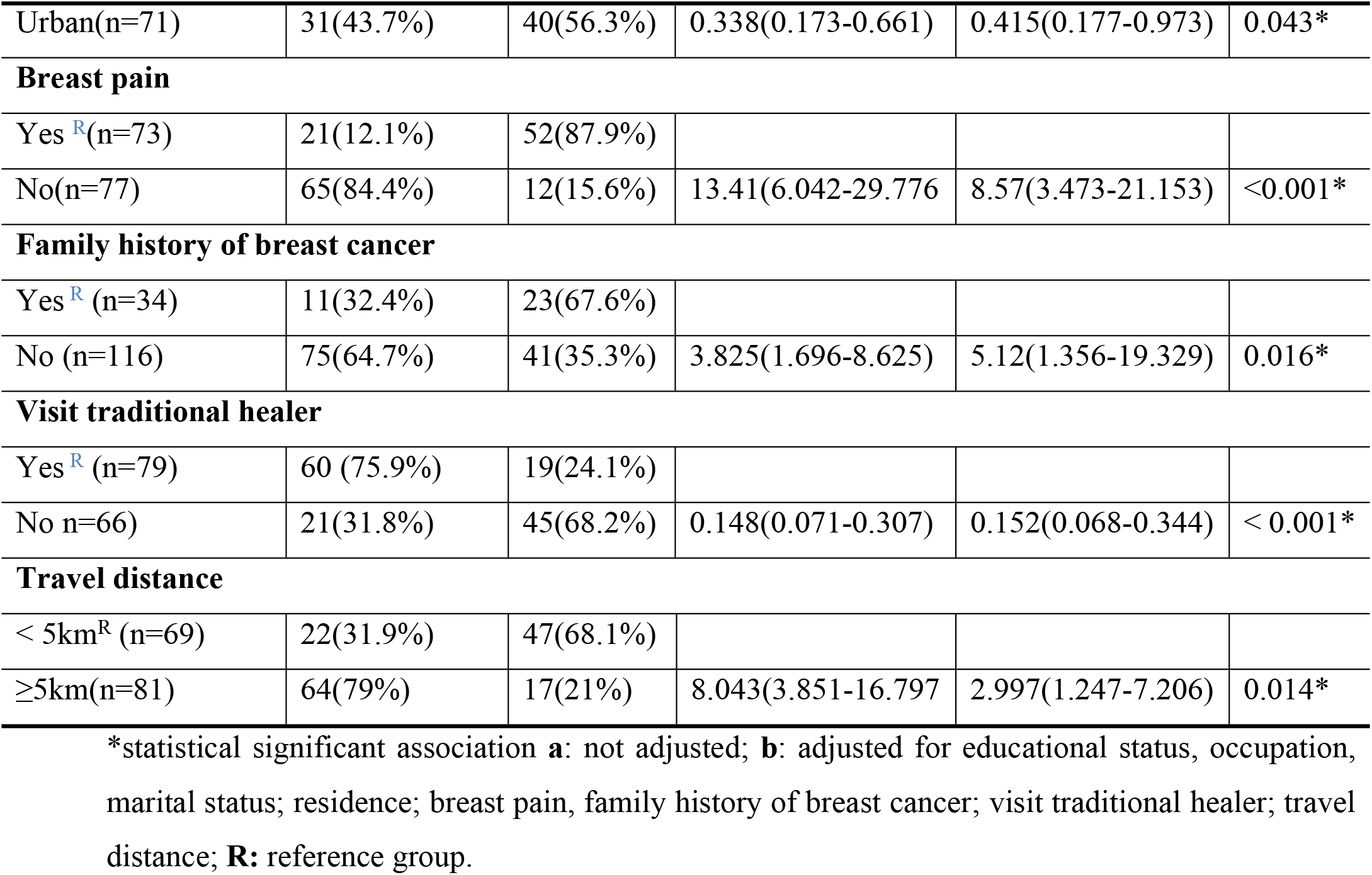
Bivariate and multivariate logistic regression indicating factors associated with delayed presentation of breast cancer patients.

## Discussion

The prevalence of patient delay in presentation has been widely reported and significantly affects BC patient survival worldwide. This study assessed delay in presentation and associated factors among BC patients at HUOC, Hawassa, Ethiopia. Our study indicated a long patient delay in presentation. The mean time to consult a health professional after notice of a first symptom prior to a BC diagnosis was 5.5 months. Patients who presented without a ≥3 months delay significantly more often received a diagnosis at an earlier stage with resulting better prognosis.

Our findings add results from Southern Ethiopia to similar studies conducted in north-central Ethiopia (14) and south and southeast Ethiopia (29) as well as from Senegal (23) and Rwanda (22). Comparable studies from Gondar and Bahir Dar, Ethiopia (15), as well as Nigeria (30) reported even longer times (8 and 7.7 months, respectively), while studies conducted in China (31) and Brazil (32) reported lower mean times. A resulting patient delay was found in 57.3% (95%CI=49-65.4%) of patients in our cohort which is higher than studies done in Iran (33), South Africa (34) and Dessie, North-central Ethiopia (14) but lower than the study in South and southeast Ethiopia (29), Senegal (23) as well as Gondar and Bahir Dar, Ethiopia (15). These differences may be due to different levels of awareness and socioeconomic differences between countries as well as Ethiopian regions. Ethiopia is a low-income country, with a lower per capita income than China or Brazil, with a generally poor health infrastructure and low awareness of people about the non-communicable diseases, women may delay in presentation to health facilities (35)(36). Moreover, the observed differences might account for variations in sample size and different cut-off points for classifying patient delay.

Generally, reduction of patient delay leads to diagnosis in earlier stages of diseases which are associated with reduced mortality (37). In our study, patients who were diagnosed without delay were significantly more often diagnosed in earlier stages (unadjusted OR=8.02). There is a need to improve community awareness on BC prevention and early detection to decrease delays in the diagnosis of BC. The Ethiopian national cancer control plan of 2016-2020, based on the world health organization’s global cancer control strategy, has been promoting steps to reducing cancer morbidity and mortality in Ethiopia since 2016. However our study among others (29) (14) (38) points the high amount of delayed diagnosis and numbers of breast cancer patients seeking care. This emphasizes the need of further improvements and the implementation of effective strategies to increase awareness and promote first signs of BC for an early detection of BC.

Our study indicated that home residence and travel distance, breast pain, family history of BC, and visit traditional healer are significantly associated with patient delay. Furthermore, we found an association between higher education and less delayed diagnoses. This might be due to women with higher educational levels having a chance to get up-to-date information, and review different sources, which would increase their knowledge of BC risks, early symptoms, and treatment options, which might increase their awareness of the disease and elevate their medical help-seeking behavior. Women who were from urban areas were less likely to delay than rural residents. Women living in rural areas may have difficulties with transportation to health facilities and oncology hospitals which are set in urban settings. The finding is consistent with the studies done in South Africa (34) and Ethiopian cities Dessie (14), Gondar, and Bahir Dar (15). In our study, patients who had not noticed breast pain were nine times more likely to delay than their counterparts. Those patients who travel more than five kilometers to the next health provider were about three times more likely to delay than their counterparts. This finding is supported by the study done in northwest Ethiopia (15). Long distance travel may induce a patient delay in seeking medical early since transportation is costly and often time-Consuming. Painful and severe symptoms are known to trigger medical help-seeking (39). Our results are consistent to results from Egypt in which women without pain symptoms presented more often with a relevant delay (13). BC patients who have not visited a traditional healer were less likely to delay – a results which is consistent with findings from Rwanda (22) and Libya (18). According to a study conducted in Harar, Eastern Ethiopia and more than 60% of households use traditional medicine as part of their health care system (38). Similarly the study done in south and southwest Ethiopia indicated that, of the 426 total patients in the study across several hospitals, 58.8% used traditional medicine (29). Another study indicated that traditional healers were most commonly visited by cancer patients (40). These visits delay early presentation in the health care system and contribute to the progression of the disease. The ministry of health should develop a solid protocol for traditional healers’ education on curative treatment options for cancer; perhaps it decreases patients’ delay in presentation.

### Strength and Limitations

The strengths of our study lies in the international comparability due to the use of the Aarhus recommendation (28). Moreover, the exceptionally high response rate lowers the risk of bias. However, the cross-sectional nature of the study limits the cause-effect relationship and recall, and social desirability bias related to not remembering the exact date of their first symptom recognition could limit the validity of the results. Nevertheless, these limitations were addressed by especially training the data collectors to minimize recall and social desirability bias among participants.

## Conclusion

This cross-sectional questionnaire-based study found a frequently long patient delay of BC patients from recognition of the first symptoms to the first presentation at a diagnostic facility as well as a high share of late-stage diagnosis among delayed patients in Hawassa, Ethiopia. Rural residence, breast pain, no family history of BC, visit of a traditional healer before diagnosis, and travel distance of >5 km were important significant factors of long patient delay. These findings emphasize the need to improve community, health care professionals, and licensed traditional healers’ awareness about the significance of early health care seeking following breast symptoms and early referral following suspicious breast abnormalities.

## Data Availability

Data are available upon reasonable request.

## Abbreviations

AOR: adjusted odds ratio
ETB: Ethiopian birr
HUCSH: Hawassa university comprehensive specialized hospital
SPSS: statistical package for social science
HUOC: Hawassa University Oncology Center
TASH: Tikur Anbessa specialized hospital.

## Acknowledgments

We would like to acknowledge Hawassa University and Else Kröner-Fresenius-Stiftung for funding the study and we are also grateful to the study participants for their cooperation.

## Declarations

### Consent For publication

Not applicable. Individuals’ information, Images, or videos are not included in this report

## Availability of data and materials

The dataset collected and analyzed in this study is available from the corresponding Author upon reasonable request.

## Author contributions

### Conceptualization

Jabir Abdella Muhammed, Eric Sven Kroeber, Bedilu Deribe, Susanne Unverzagt, Lesley Taylor

### Data curation

Jabir Abdella Muhammed, Eric Sven Kroeber, Susanne Unverzagt, Amdehiwot Aynalem, Deriba Fetene

### Formal analysis

Jabir Abdella Muhammed, Eric Sven Kroeber, Susanne Unverzagt

### Funding acquisition

Jabir Abdella Muhammed

### Methodology

Jabir Abdella Muhammed, Eric Sven Kroeber, Bedilu Deribe, Susanne Unverzagt, Lesley Taylor, Amdehiwot Aynalem

### Project administration

Jabir Abdella Muhammed, Bedilu Deribe, Lesley Taylor

### Software

Jabir Abdella Muhammed

### Supervision

Bedilu Deribe, Lesley Taylor

### Validation

Eric Sven Kroeber, Bedilu Deribe, Susanne Unverzagt, Lesley Taylor,Deriba fetene

### Writing-original draft

Jabir Abdella Muhammed

### Writing-review and editing

Jabir Abdella Muhammed, Eric Sven Kroeber, Bedilu Deribe, Susanne Unverzagt, Lesley Taylor, Amdehiwot Aynalem, Deriba fetene

## Notes

### Competing Interest Statement

The authors have declared no competing interest.

### Funding Statement

this fund was supported by Else Kröner-Fresenius-Stiftung. The funder had no role in study design, data collection. Grant number is not applicable

